# Basic determinants of child growth in sub-Saharan Africa: cross-sectional survey analysis of positive deviants in poor households

**DOI:** 10.1101/19006759

**Authors:** Dickson A. Amugsi, Zacharie T. Dimbuene

## Abstract

**Background:** Childhood malnutrition is a significant public health problem confronting countries across the globe. Nonetheless, recent evidence suggests a downward trend in undernutrition among children globally. Despite the progress made at the global level, sub-Saharan Africa did not experience significant improvement in the past decades. The objective of this study was to investigate the basic determinants associated with linear growth among children under 5 years living in poor households.

**Methods:** The study used nationally representative cross-sectional survey data from Ghana, Kenya, Democratic Republic of Congo (DRC), Nigeria and Mozambique. The participants are children aged 0–59 months (N=24,264) living in poor households. The DHS obtained information on children through face-to-face interviews with mothers. The height of the children was also measured and used to compute the height-for-age Z-scores (HAZ). In this study, HAZ is categorised into HAZ>-2 standard deviations (SD) (not stunted/better growth) and HAZ<-2 SD (stunted/poor growth).

**Results:** A unit change in maternal years of education was associated with increased odds of better growth among children living in poor households in DRC [adjusted odds ratio (aOR)= 1.03, 95% CI=1.01,1.07)], Ghana (aOR=1.06, 95% CI=1.01,1.11), Kenya (aOR=1.03, 95% CI= 1.01, 1.05) and Nigeria (aOR=1.08, 95%=1.06,1.10). Maternal antenatal attendance of at least four visits was associated positively with better child growth in DRC (aOR=1.32, 95% CI=1.05, 1.67) and Ghana (aOR=1.67, 95% CI=1.19, 2.33). In Ghana, Mozambique and DRC, breastfeeding was associated significantly with the likelihood of better linear growth when only socio-demographic correlates were included in the models but disappeared after the inclusion of child-level covariates. In Nigeria, normal maternal weight was associated with increased odds (aOR=1.24, 95% CI=1.08, 1.43) of positive growth among children living in poor households, so was overweight (aOR=1.51, 95% CI= 1.24, 1.83). In all the countries except Ghana, child biological factors such as sex and age were associated with reduced odds of better growth.

**Conclusions:** The socio-demographic factors included in this analysis have the potential to promote linear growth of children under 5 years living in poor households. Interventions aimed at fostering linear growth among children living in poverty should target at enhancing these factors.

## Introduction

Childhood undernutrition is a significant public health problem confronting countries across the globe. However, there is evidence that global trends in child malnutrition have improved over the years. For example, it is estimated that childhood stunting (short stature for age), a critical undernutrition metric, decreased from 39·7% in 1990 to 26·7% in 2010 (1). The trend is predicted to reduce to 22% in 2020 (1). Despite global-level progress in the reduction of the problem during the past decades (2), Africa has not seen much improvement. There was a decline in stunting trends from 40.5% in 1980 to 35.2% in 2000 (3), the trends stagnated at 40% between 1990 and 2010 (1). The level of decline in sub-Saharan Africa (SSA) is not different from the African region as a whole. For example, in SSA, the stunting trends decreased from 43% in 2000 to 34% in 2018 (4). The risk factors contributing to the high stunting prevalence in Africa are well documented (5-9).

The consequences of stunting on the later life of the child are well known. Indeed, there is strong evidence that stunting can have long-term effects on cognitive development, school achievement, economic productivity in adulthood and maternal reproductive outcomes (10-13). Stunting is also a condition that may be very difficult to reverse (10). Given the negative consequences of stunting on child health outcomes, the international community has paid considerable attention to the problem. For instance, the World Health Assembly (WHA) Resolution (2012) set a 40% reduction in the number of stunted children under-5 as one of the six global nutrition targets for 2025 (11, 14). The Sustainable Development Goals (SDGs) (15) also captured this undernutrition metric as a critical developmental target. The above discussion suggests the need for substantial investment in nutrition interventions to address childhood stunting, as averting stunting could produce life-long benefits. However, to achieve this goal, a clear understanding of the broader factors that promote child growth is necessary to provide evidence for the design of effective nutrition interventions. The present study is set out to provide this evidence by focusing on factors that foster child growth rather than risk factors of child growth deficiencies.

The evidence further suggests a disproportionate burden of stunting among children in low and middle-income countries, which is attributable to poverty, lack of food and high incidence of infectious diseases among others (3, 11, 16, 17). In sub-Saharan Africa (SSA), several countries are confronted with a high prevalence of stunting among children under five years of age (3, 11, 17). The problem is notably more severe among children living in poor households (17-19)—they tend to have the highest prevalence of childhood stunting (17). It is the case because poverty creates conditions that favour poor child growth outcomes and prevents affected populations from obtaining adequate access to prevention and care (17). Despite the health challenges facing children living in poverty, some children are living in the same conditions *(positive deviants)* or even worse yet have positive growth outcomes comparable to children residing in privilege households anywhere in the world (20-23). The questions this paper intends to address is why some children in poor households are not stunted, although they are faced with similar adversity as those who are stunted are? What are the possible factors that help them to have better growth outcomes? Understanding this will help design programmes to promote the growth of children in impoverished households or environments.

The concept of *positive deviance* (as referenced above) is based on the premise is that even in places where deprivation is severe and widespread, some families are able to cope and harness scant resources, sufficient to support optimal child development *(24, 25)*. The positive deviance (PD) approach is founded on idea that problems can be overcome using solutions that already exist within the community (26, 27). PD often studies the behaviours and characteristics of individuals who have better health outcomes than their peers who live in the same community (26). The PD approach was used previously to investigate several health-related issues in diverse settings (28-36). In statistical analysis, it is often quantified as those who do not experience a negative outcome of interest compared to those around them with the same resources (26). Using the PD approach can be useful because it studies the ‘positive’ aspects of an outcome or community instead of the ‘negative’, and can identify potential points of intervention. The *positive deviants* in the present study are children who live in poor households and yet are not stunted relative to their counterparts who live in the same environment but are stunted. The objective of the present study is to examine the basic factors associated with better growth outcomes among children living in poor households. This resource-focused approach moves away from the dominant risk model approach, where the focus is usually on risk factors of child growth deficiencies. Using the PD approach is to help understand the drivers of better child growth and interventions to promote these drivers in poor households effectively.

## Methodology

### Data sources and sampling strategy

We analysed the Demographic and Health Surveys (DHS) (37) data from Ghana, Kenya, Nigeria, Mozambique and Democratic Republic of Congo (DRC) (38, 39). The DHS are nationally representative surveys conducted every five years in lower- and middle-income countries, using the same questionnaires to enable comparison across countries (40, 41). The DHS utilises a two-stage sample design. The detail description of the DHS design and sampling strategies can be found elsewhere (42-46). The DHS data collectors then interviewed all eligible study participants in their respective households of each country using standardised questionnaires and interview protocols. In this analysis, we used data of children aged 0–59 months and their mothers aged 15-49 years. The DHS obtained children data through face-to-face interviews with their mothers or caregivers. The length/height of the children was measured using an adjustable measuring board calibrated in millimeters. Recumbent length (lying down on the board) was measured for young children while standing height was measured for older children. The height data were converted into Z-scores based on the 2006 WHO growth standards, taking into account the age and sex of the child (47). The total samples used in the present analysis were: Ghana, n= 1,453; Nigeria, n= 10,378; Kenya, n= 4,967; Mozambique, n= 3,487; and DRC, n= 3,979.

## Ethics statement

The government recognized Ethical Review Committees of the respective countries approved the DHS study protocols. Besides, the ethical clearance was granted by the Institutional Review Board of ICF International, USA before the surveys were conducted. Informed consent was obtained from the mothers of the study children before they were included in the study. The DHS Program permitted the authors to use the data. The data were wholly anonymised, and therefore, the authors did not seek further ethical clearance before their use.

### Outcome and predictor variables

#### Outcome Variables

We used the child height-for-age Z-scores (HAZ) as the indicator of child linear growth in the analysis. For all datasets, HAZ scores were computed using 2006 WHO growth standards (47). We reclassified the child HAZ into stunted (poor growth) and not stunted (better growth). Children who have HAZ above –2 SD (HAZ>-2SD) (47, 48) were considered having a better linear growth and described in this study as positive deviants. Similarly, children who have HAZ below –2 SD (HAZ<-2) from the median HAZ of the WHO reference population (47) were considered stunted or having poor growth. It is significant to underscore that DHS data contained all the three indicators of child nutritional status: height-for-age z-scores (HAZ), weight-for-age z-scores (WAZ) and weight-for-height z-scores (WHZ). However, we opted for HAZ because it is a cumulative indicator of a child nutritional status, and therefore more informative and appropriate for use in the positive deviance analysis. The WHZ, on the other hand, reflects more recent processes often associated with acute food shortages and/or illnesses leading to weight loss, while the WAZ lies between HAZ and WHZ. For example, a child who has poor HAZ is also likely to be underweight, so is a child who has poor WHZ.

We stratified the analysis by household wealth index (WI). The WI has been used severally as an indicator for measuring inequalities associated with health outcomes as well as expenditure and income among households (41, 42, 44, 49). The detail discussion on how the DHS created the WI is well documented (40, 41, 44). In the datasets, the WI is classified into five quintiles: poorest, poor, middle, richer and richest(40, 41, 44). In this paper, we recoded poor and poorest into poor/worse-off households. We combined the poorest and poor households wealth quintiles because the literature suggests that children in these households have similar health outcomes (17-19). We restricted all the analyses to children living in these households.

#### Analytical framework

The modified UNICEF conceptual framework underpinned our analysis (50, 51). This framework outlines how the various factors/determinants influence child survival, growth and development at different levels. These factors are analysed in terms of immediate, underlying and basic determinants. The immediate determinants are adequate nutrients intake and health, while the underlying determinants are food security, care for children and women, healthcare and a healthy environment (51). The underlying determinants either influence child health directly or through the immediate determinants. The basic determinants, in turn, influence the underlying determinants. In this context, the basic determinants are described as “exogenous” factors, which effects on child nutrition are through their effects on the intervening proximate/underlying determinants. Thus, the underlying determinants are endogenously determined by the exogenous determinants (52). For example, the effect of an exogenous variable such as maternal education on child growth outcomes is through its impact on good child-caring practices, including high utilisation of health care services, among others by educated mothers.

## Data analysis

The present empirical analysis focused mainly on the basic determinants (i.e. socio-demographic factors). The scientific basis for this type of analysis is well documented (52-55). Apart from the socio-demographic factors, variables such as antenatal care (ANC) and breastfeeding practices, which depend mostly on exogenous public health provisions (52) were included in our empirical models. The significance for the inclusion these two variables is that improvement or otherwise of ANC and breastfeeding practices are more likely to inform policies, programmes and interventions rather than changes in socio-demographic endowments of households (52). For example, the available data suggest that policy, institutional and contextual settings are critical determinants of the prevalence of breastfeeding practices (52, 56). In the analysis, we built two regression models for each of the five countries. In the first model, we included the various socio-demographic factors [maternal body mass index (BMI), education, age, work status, parity, breastfeeding practices, marital status, ANC, the gender of head fo household, size of household, total children under five years and place of residence]. We adjusted for child dietary diversity (DD)—the details of how the DD is created can be found elsewhere (38), age and sex in the second and final model. The conceptual framework and the literature guided the selection of the explanatory variables (51). We estimated adjusted odds ratio (aORs) of the effects of the basic determinants on child growth in poor households.

## Results

### Characteristics of study samples

Tables 1 presents the results of the descriptive analysis. The results showed that Ghana (76%) has the highest number of children with better growth followed by Kenya (68%), while in Mozambique, DRC and Nigeria, the prevalence ranged from 50% to 52%. Regarding dietary diversity intake, Mozambique had the highest prevalence of children who consumed at least four food groups (24%), with DRC (6%) and Nigeria (6%) having the lowest prevalence. Similarly, Mozambique had the highest number of women with normal weight (85%). The prevalence ranged from 68% to 76% in DRC, Ghana, Kenya and Nigeria. For maternal education, Ghana has the highest prevalence (23%) of women who had attained a secondary school education, while Mozambique has the lowest prevalence (1.20%). Higher education was less than 1% among women in poor households across all countries. Regarding antenatal attendance among women, DRC registered the highest prevalence (77%) followed by Ghana (59%), while Nigeria registered the lowest prevalence (19%).

**Table 1:**
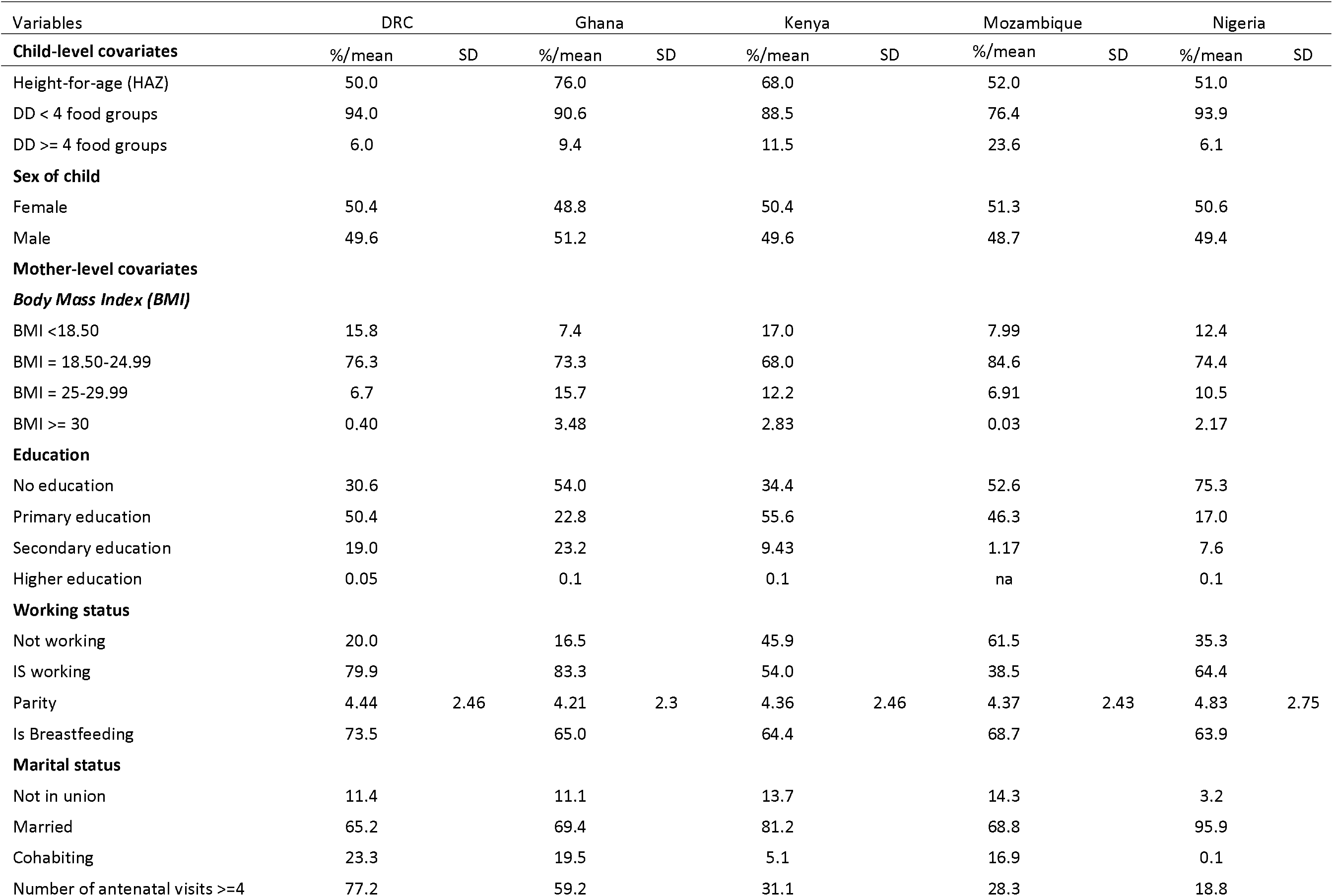
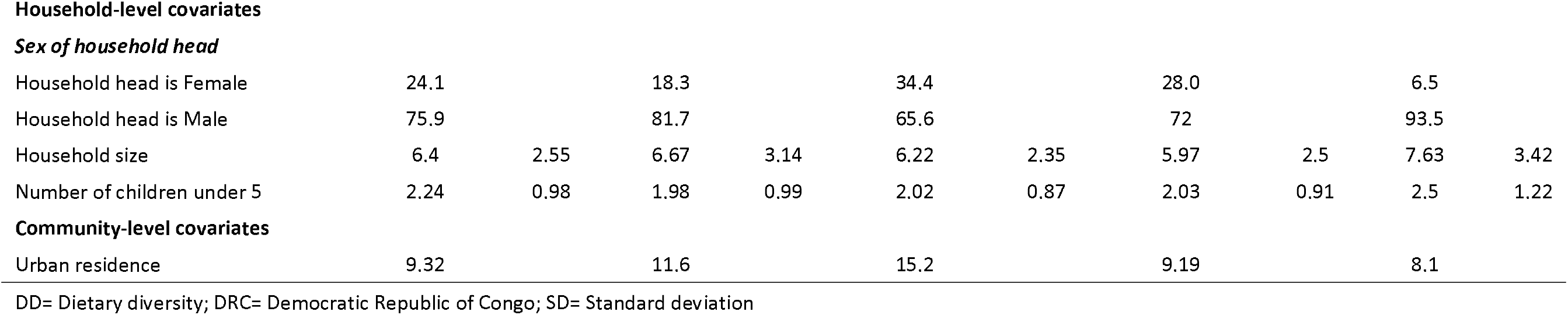
Characteristics of the study samples of the five countries.

| Variables | DRC |  | Ghana |  | Kenya |  | Mozambique |  | Nigeria |  |
| --- | --- | --- | --- | --- | --- | --- | --- | --- | --- | --- |
| **Child-level covariates** | %/mean | SD | %/mean | SD | %/mean | SD | %/mean | SD | %/mean | SD |
| Height-for-age (HAZ) | 50.0 |  | 76.0 |  | 68.0 |  | 52.0 |  | 51.0 |  |
| DD < 4 food groups | 94.0 |  | 90.6 |  | 88.5 |  | 76.4 |  | 93.9 |  |
| DD >= 4 food groups | 6.0 |  | 9.4 |  | 11.5 |  | 23.6 |  | 6.1 |  |
| **Sex of child** |  |  |  |  |  |  |  |  |  |  |
| Female | 50.4 |  | 48.8 |  | 50.4 |  | 51.3 |  | 50.6 |  |
| Male | 49.6 |  | 51.2 |  | 49.6 |  | 48.7 |  | 49.4 |  |
| **Mother-level covariates** |  |  |  |  |  |  |  |  |  |  |
| ***Body Mass Index (BMI)*** |  |  |  |  |  |  |  |  |  |  |
| BMI <18.50 | 15.8 |  | 7.4 |  | 17.0 |  | 7.99 |  | 12.4 |  |
| BMI = 18.50-24.99 | 76.3 |  | 73.3 |  | 68.0 |  | 84.6 |  | 74.4 |  |
| BMI = 25-29.99 | 6.7 |  | 15.7 |  | 12.2 |  | 6.91 |  | 10.5 |  |
| BMI >= 30 | 0.40 |  | 3.48 |  | 2.83 |  | 0.03 |  | 2.17 |  |
| **Education** |  |  |  |  |  |  |  |  |  |  |
| No education | 30.6 |  | 54.0 |  | 34.4 |  | 52.6 |  | 75.3 |  |
| Primary education | 50.4 |  | 22.8 |  | 55.6 |  | 46.3 |  | 17.0 |  |
| Secondary education | 19.0 |  | 23.2 |  | 9.43 |  | 1.17 |  | 7.6 |  |
| Higher education | 0.05 |  | 0.1 |  | 0.1 |  | na |  | 0.1 |  |
| **Working status** |  |  |  |  |  |  |  |  |  |  |
| Not working | 20.0 |  | 16.5 |  | 45.9 |  | 61.5 |  | 35.3 |  |
| Is working | 79.9 |  | 83.3 |  | 54.0 |  | 38.5 |  | 64.4 |  |
| Parity | 4.44 | 2.46 | 4.21 | 2.3 | 4.36 | 2.46 | 4.37 | 2.43 | 4.83 | 2.75 |
| Is Breastfeeding | 73.5 |  | 65.0 |  | 64.4 |  | 68.7 |  | 63.9 |  |
| **Marital status** |  |  |  |  |  |  |  |  |  |  |
| Not in union | 11.4 |  | 11.1 |  | 13.7 |  | 14.3 |  | 3.2 |  |
| Married | 65.2 |  | 69.4 |  | 81.2 |  | 68.8 |  | 95.9 |  |
| Cohabiting | 23.3 |  | 19.5 |  | 5.1 |  | 16.9 |  | 0.1 |  |
| Number of antenatal visits >=4 | 77.2 |  | 59.2 |  | 31.1 |  | 28.3 |  | 18.8 |  |
| **Household-level covariates*****Sex of household head*** |  |  |  |  |  |  |  |  |  |  |
| Household head is Female | 24.1 |  | 18.3 |  | 34.4 |  | 28.0 |  | 6.5 |  |
| Household head is Male | 75.9 |  | 81.7 |  | 65.6 |  | 72 |  | 93.5 |  |
| Household size | 6.4 | 2.55 | 6.67 | 3.14 | 6.22 | 2.35 | 5.97 | 2.5 | 7.63 | 3.42 |
| Number of children under 5 | 2.24 | 0.98 | 1.98 | 0.99 | 2.02 | 0.87 | 2.03 | 0.91 | 2.5 | 1.22 |

**Community-level covariates**
|  |  |  |  |  |  |  |  |  |  |
| --- | --- | --- | --- | --- | --- | --- | --- | --- | --- |
| Urban residence | 9.32 |  | 11.6 |  | 15.2 |  | 9.19 |  | 8.1 |

DD= Dietary diversity; DRC= Democratic Republic of Congo; SD= Standard deviation

### Multivariable results of the association between socio-demographic factors and better child growth

Tables 2-6 present the results of the association between sociodemographic factors at the child-level, maternal-level, household-level and community-level, and better linear growth among children in five SSA countries. The results showed that a unit change in maternal years of education was associated with increased odds of better linear growth among children in DRC (aOR=1.03, 95% CI=1.01,1.07), Ghana (aOR=1.06, 95% CI=1.01,1.11), Kenya (aOR=1.03, 95% CI= 1.01, 1.05) and Nigeria (aOR=1.08, 95%=1.06,1.10). Antenatal attendance of at least four visits was associated significantly with the likelihood of better child growth in DRC (aOR=1.32, 95% CI=1.05, 1.67) and Ghana (aOR=1.67, 95% CI=1.19, 2.33). The association did not reach statistical significance in the remaining three countries. In Kenya, children of mothers who were working and live in poor households had 23% reduced odds of better growth (aOR= 0.77, 95% CI=0.66, 0.91) relative to children of non-working mothers. In Nigeria, Mozambique and DRC, breastfeeding was positively associated with better child growth, but this association disappeared after the child level covariates were included in the model. Urban place of residence was associated with 28% reduced odds of better child growth (aOR=0.72, 95% CI=0.55, 0.95) in Mozambique, and increased odds in Nigeria (aOR=1.58, 95% CI= 1.33, 1.87). In Nigeria, normal maternal weight (BMI) was associated with increased odds (aOR=1.24, 95% CI=1.08, 1.43) of better child growth. Maternal overweight was also associated with increased odds (aOR=1.51, 95% CI= 1.24, 1.83) of better child growth in Nigeria. A unit change in household size was associated with increased odds (aOR=1.05, 95% CI= 1.01, 1.10) of better child growth. Maternal parity reduces the odds of better child growth (aOR=0.95, 95% CI=0.92, 0.98) in Nigeria. In all the countries except Ghana, child biological factors such as sex and age were associated with reduced odds of better child growth.

**Table 2:** Multivariable analysis of the effects of socio-demographic factors on better linear growth among children living in poor households in DRC.

| Variables | Model 1 | Model 2 |
| --- | --- | --- |
| **Mother-level covariates** |  |  |
| BMI (kg/m<sup>2</sup>) = 18.50 - 24.99 | 0.972<br>(0.758 - 1.247) | 0.979<br>(0.753 - 1.274) |
| BMI (kg/m<sup>2</sup>) = 25 - 29.99 | 0.913<br>(0.604 - 1.379) | 0.861<br>(0.551 - 1.348) |
| BMI (kg/m<sup>2</sup>) >= 30 | 0.310*<br>(0.0795 - 1.207) | 0.301<br>(0.0558 - 1.620) |
| Maternal education (in single years) | 1.030**<br>(1.001 - 1.060) | 1.034**<br>(1.003 - 1.065) |
| Age of the mother (in years) | 0.994<br>(0.972 - 1.016) | 1.012<br>(0.988 - 1.037) |
| Working status = Is working | 0.840<br>(0.671 - 1.052) | 0.873<br>(0.684 - 1.113) |
| Parity | 0.995<br>(0.934 - 1.060) | 0.989<br>(0.923 - 1.061) |
| Is Breastfeeding = YES | 1.379***<br>(1.110 - 1.712) | 0.813*<br>(0.637 - 1.036) |
| Marital Status = Married | 0.880<br>(0.648 - 1.196) | 0.937<br>(0.677 - 1.297) |
| Marital Status = Cohabiting | 0.986<br>(0.704 - 1.379) | 1.036<br>(0.731 - 1.469) |
| Number of antenatal visits = 4+ visits | 2.125***<br>(1.710 - 2.641) | 1.321**<br>(1.046 - 1.668) |
| **Household-level covariates** |  |  |
| Head of HH is Male | 0.957<br>(0.759 - 1.206) | 0.936<br>(0.741 - 1.183) |
| Household size | 1.014<br>(0.965 - 1.065) | 1.005<br>(0.954 - 1.058) |
| Number of children under 5 years | 1.018<br>(0.904 - 1.147) | 1.068<br>(0.943 - 1.210) |
| **Community-level covariates** |  |  |
| Urban residence = Urban | 0.989<br>(0.740 - 1.322) | 0.972<br>(0.720 - 1.312) |
| **Child-level covariates** |  |  |
| Dietary Diversity (DD) >= 4 |  | 1.049<br>(0.686 - 1.602) |
| Age of the child (in months) |  | 0.959***<br>(0.953 - 0.965) |
| Sex of child = Male |  | 0.773***<br>(0.639 - 0.935) |
| **Observations** | **3,979** | **3,979** |

95% Confidence Intervals (CIs) in parentheses; DD-Dietary diversity; HH-Household; BMI-Body mass index
\*\*\* p<0.01, \*\* p<0.05, \* p<0.1

**Table 3:** Multivariable analysis of the effects of socio-demographic factors on better linear growth among children living in poor households in Ghana.

| Variables | Model 1 | Model 2 |
| --- | --- | --- |
| **Mother-level covariates** |  |  |
| BMI (kg/m<sup>2</sup>) = 18.50 - 24.99 | 0.944<br>(0.554 - 1.608) | 0.946<br>(0.554 - 1.617) |
| BMI (kg/m<sup>2</sup>) = 25 - 29.99 | 1.702<br>(0.888 - 3.262) | 1.727*<br>(0.902 - 3.305) |
| BMI (kg/m<sup>2</sup>) >= 30 | 2.048<br>(0.739 - 5.677) | 2.183<br>(0.783 - 6.089) |
| Maternal education (in single years) | 1.059**<br>(1.012 - 1.107) | 1.057**<br>(1.010 - 1.106) |
| Age of the mother (in years) | 1.004<br>(0.966 - 1.044) | 1.009<br>(0.969 - 1.050) |
| Working status = Is working | 0.772<br>(0.509 - 1.170) | 0.798<br>(0.524 - 1.214) |
| Parity | 1.009<br>(0.902 - 1.129) | 1.007<br>(0.899 - 1.129) |
| Is Breastfeeding = YES | 1.705***<br>(1.206 - 2.410) | 1.412*<br>(0.975 - 2.045) |
| Marital Status = Married | 1.316<br>(0.761 - 2.278) | 1.300<br>(0.755 - 2.239) |
| Marital Status = Cohabiting | 0.994<br>(0.539 - 1.831) | 0.950<br>(0.516 - 1.747) |
| Number of antenatal visits = 4+ visits | 2.004***<br>(1.464 - 2.743) | 1.667***<br>(1.193 - 2.329) |
| **Household-level covariates** |  |  |
| Head of HH is Male | 0.889<br>(0.549 - 1.440) | 0.911<br>(0.566 - 1.468) |
| Household size | 0.987<br>(0.928 - 1.050) | 0.983<br>(0.923 - 1.047) |
| Number of children under 5 | 0.946<br>(0.774 - 1.156) | 0.942<br>(0.771 - 1.150) |
| **Community-level covariate** |  |  |
| Urban residence = Urban | 1.239<br>(0.735 - 2.087) | 1.224<br>(0.733 - 2.046) |
| **Child-level covariates** |  |  |
| Dietary Diversity (DD) >= 4 |  | 1.281<br>(0.765 - 2.146) |
| Age of the child (in months) |  | 0.989*<br>(0.979 - 1.000) |
| Sex of child = Male |  | 0.850<br>(0.624 - 1.159) |
| **Observations** | **1,453** | **1,453** |

95% Confidence Intervals (CIs) in parentheses; DD-Dietary diversity; HH-Household; BMI-Body mass index
\*\*\* p<0.01, \*\* p<0.05, \* p<0.1

**Table 4:**
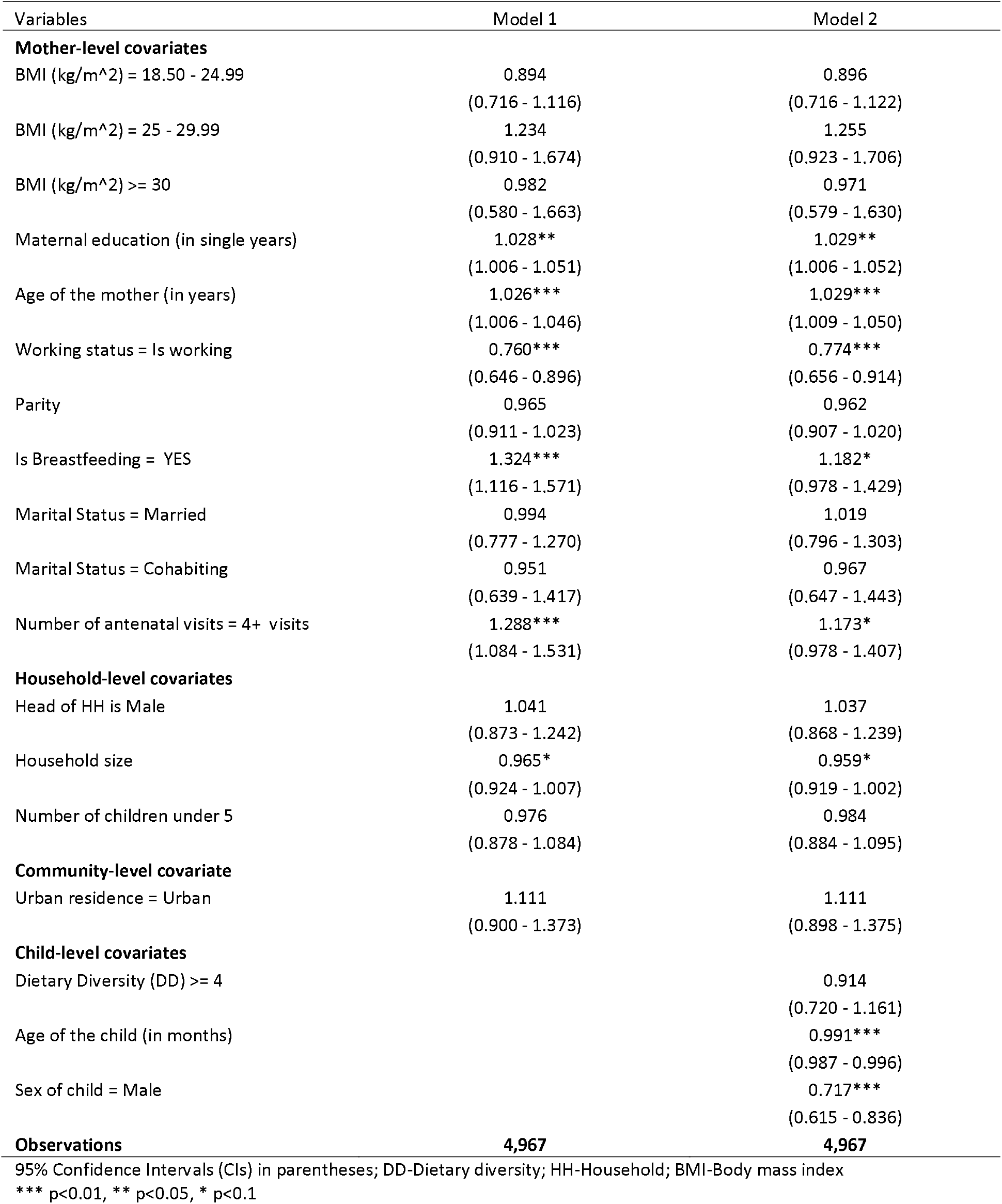
Multivariable analysis of the effects of socio-demographic factors on better linear growth among children living in poor households in Kenya.

| Variables | Model 1 | Model 2 |
| --- | --- | --- |
| **Mother-level covariates** |  |  |
| BMI (kg/m<sup>2</sup>) = 18.50 - 24.99 | 0.894<br>(0.716 - 1.116) | 0.896<br>(0.716 - 1.122) |
| BMI (kg/m<sup>2</sup>) = 25 - 29.99 | 1.234<br>(0.910 - 1.674) | 1.255<br>(0.923 - 1.706) |
| BMI (kg/m<sup>2</sup>) >= 30 | 0.982<br>(0.580 - 1.663) | 0.971<br>(0.579 - 1.630) |
| Maternal education (in single years) | 1.028**<br>(1.006 - 1.051) | 1.029**<br>(1.006 - 1.052) |
| Age of the mother (in years) | 1.026***<br>(1.006 - 1.046) | 1.029***<br>(1.009 - 1.050) |
| Working status = Is working | 0.760***<br>(0.646 - 0.896) | 0.774***<br>(0.656 - 0.914) |
| Parity | 0.965<br>(0.911 - 1.023) | 0.962<br>(0.907 - 1.020) |
| Is Breastfeeding = YES | 1.324***<br>(1.116 - 1.571) | 1.182*<br>(0.978 - 1.429) |
| Marital Status = Married | 0.994<br>(0.777 - 1.270) | 1.019<br>(0.796 - 1.303) |
| Marital Status = Cohabiting | 0.951<br>(0.639 - 1.417) | 0.967<br>(0.647 - 1.443) |
| Number of antenatal visits = 4+ visits | 1.288***<br>(1.084 - 1.531) | 1.173*<br>(0.978 - 1.407) |
| **Household-level covariates** |  |  |
| Head of HH is Male | 1.041<br>(0.873 - 1.242) | 1.037<br>(0.868 - 1.239) |
| Household size | 0.965*<br>(0.924 - 1.007) | 0.959*<br>(0.919 - 1.002) |
| Number of children under 5 | 0.976<br>(0.878 - 1.084) | 0.984<br>(0.884 - 1.095) |
| **Community-level covariate** |  |  |
| Urban residence = Urban | 1.111<br>(0.900 - 1.373) | 1.111<br>(0.898 - 1.375) |
| **Child-level covariates** |  |  |
| Dietary Diversity (DD) >= 4 |  | 0.914<br>(0.720 - 1.161) |
| Age of the child (in months) |  | 0.991***<br>(0.987 - 0.996) |
| Sex of child = Male |  | 0.717***<br>(0.615 - 0.836) |
| **Observations** | **4,967** | **4,967** |

95% Confidence Intervals (CIs) in parentheses; DD-Dietary diversity; HH-Household; BMI-Body mass index
\*\*\* p<0.01, \*\* p<0.05, \* p<0.1

**Table 5:**
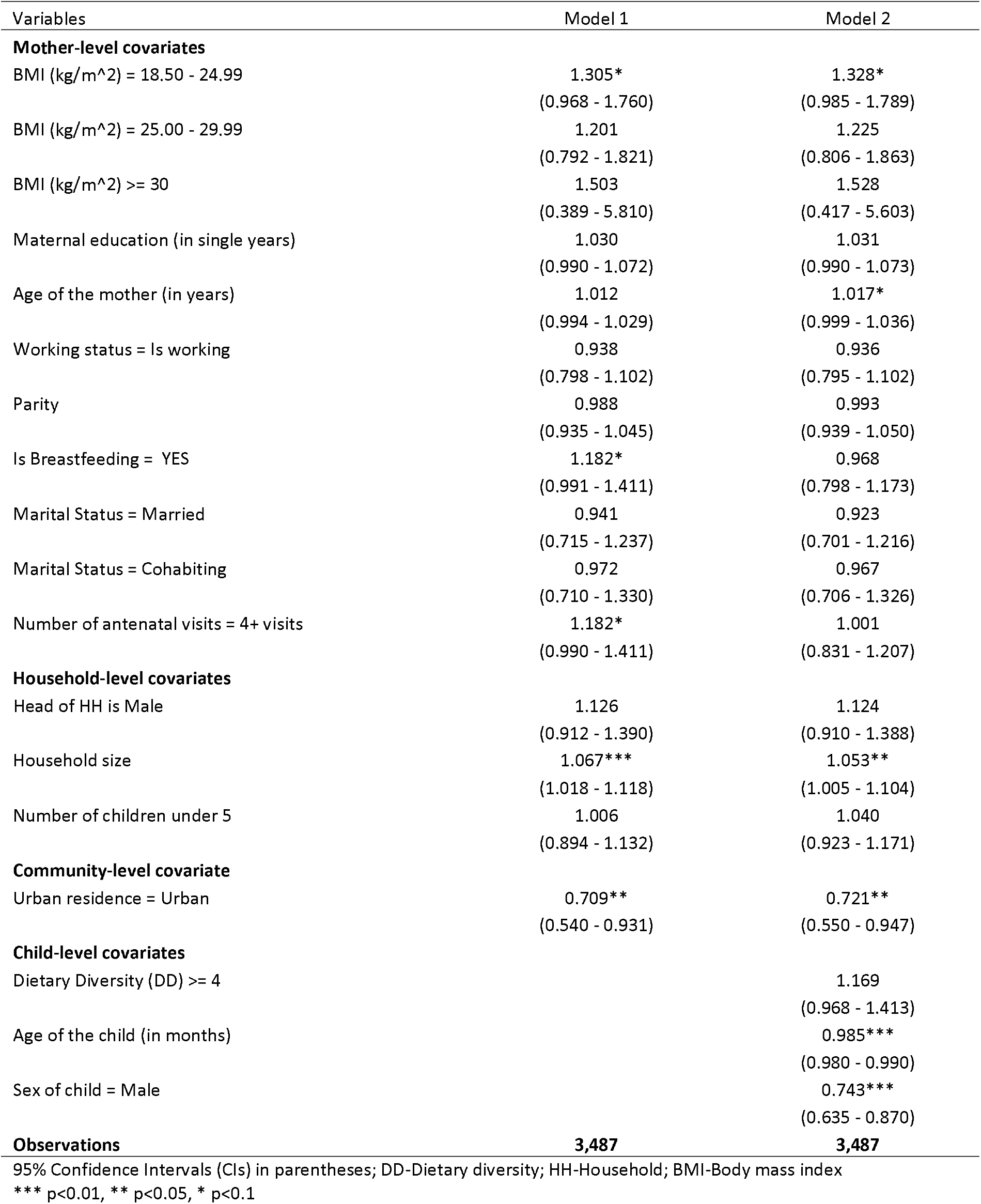
Multivariable analysis of the effects of socio-demographic factors on better linear growth among children living in poor households in Mozambique.

**Table 6:**
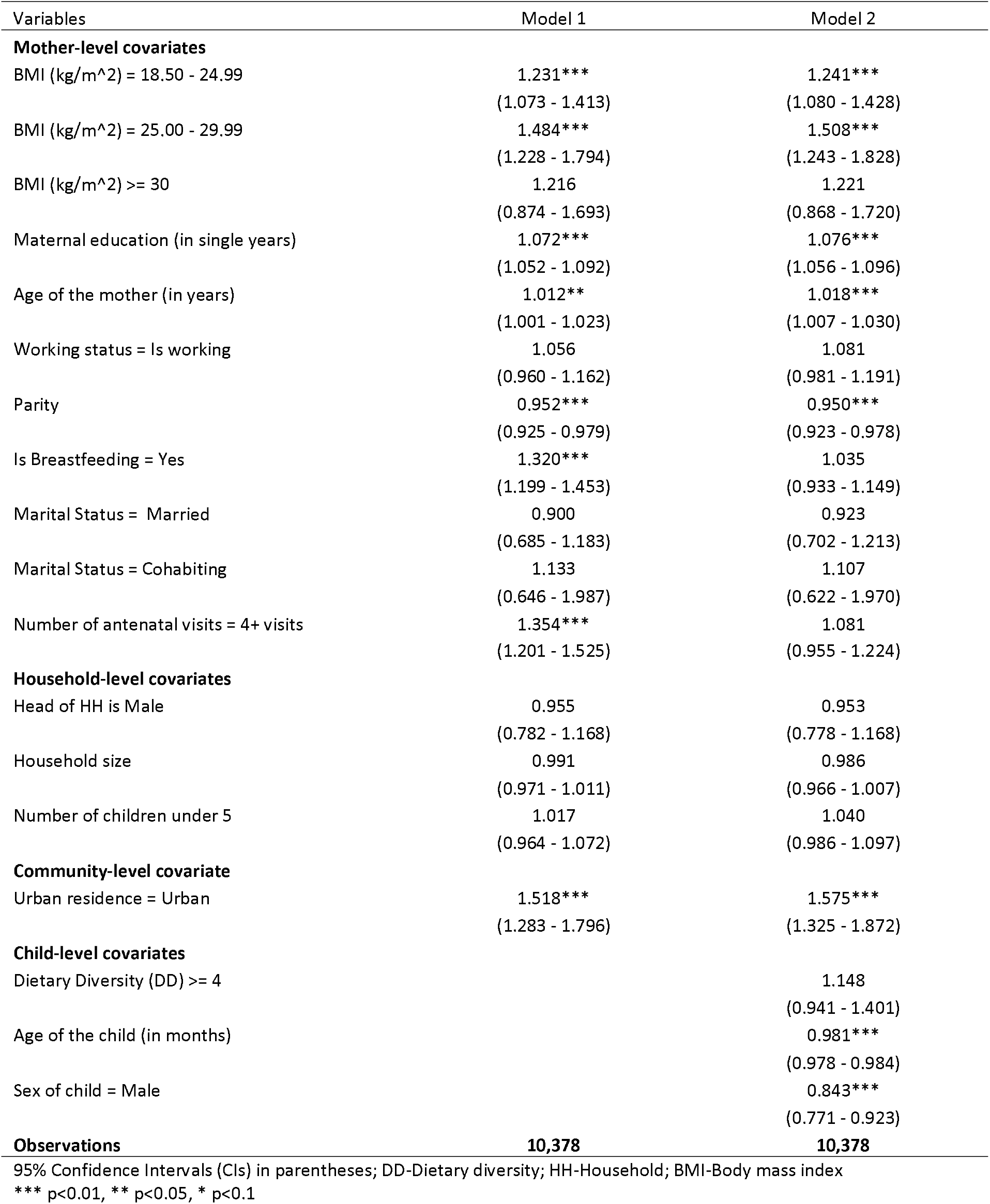
Multivariable analysis of the effects of socio-demographic factors on better linear growth among children living in poor households in Nigeria.

| Variables | Model 1 | Model 2 |
| --- | --- | --- |
| **Mother-level covariates** |  |  |
| BMI (kg/m<sup>2</sup>) = 18.50 - 24.99 | 1.231***<br>(1.073 - 1.413) | 1.241***<br>(1.080 - 1.428) |
| BMI (kg/m<sup>2</sup>) = 25.00 - 29.99 | 1.484***<br>(1.228 - 1.794) | 1.508***<br>(1.243 - 1.828) |
| BMI (kg/m<sup>2</sup>) >= 30 | 1.216<br>(0.874 - 1.693) | 1.221<br>(0.868 - 1.720) |
| Maternal education (in single years) | 1.072***<br>(1.052 - 1.092) | 1.076***<br>(1.056 - 1.096) |
| Age of the mother (in years) | 1.012**<br>(1.001 - 1.023) | 1.018***<br>(1.007 - 1.030) |
| Working status = Is working | 1.056<br>(0.960 - 1.162) | 1.081<br>(0.981 - 1.191) |
| Parity | 0.952***<br>(0.925 - 0.979) | 0.950***<br>(0.923 - 0.978) |
| Is Breastfeeding = Yes | 1.320***<br>(1.199 - 1.453) | 1.035<br>(0.933 - 1.149) |
| Marital Status = Married | 0.900<br>(0.685 - 1.183) | 0.923<br>(0.702 - 1.213) |
| Marital Status = Cohabiting | 1.133<br>(0.646 - 1.987) | 1.107<br>(0.622 - 1.970) |
| Number of antenatal visits = 4+ visits | 1.354***<br>(1.201 - 1.525) | 1.081<br>(0.955 - 1.224) |
| **Household-level covariates** |  |  |
| Head of HH is Male | 0.955<br>(0.782 - 1.168) | 0.953<br>(0.778 - 1.168) |
| Household size | 0.991<br>(0.971 - 1.011) | 0.986<br>(0.966 - 1.007) |
| Number of children under 5 | 1.017<br>(0.964 - 1.072) | 1.040<br>(0.986 - 1.097) |
| **Community-level covariate** |  |  |
| Urban residence = Urban | 1.518***<br>(1.283 - 1.796) | 1.575***<br>(1.325 - 1.872) |
| **Child-level covariates** |  |  |
| Dietary Diversity (DD) >= 4 |  | 1.148<br>(0.941 - 1.401) |
| Age of the child (in months) |  | 0.981***<br>(0.978 - 0.984) |
| Sex of child = Male |  | 0.843***<br>(0.771 - 0.923) |
| **Observations** | **10,378** | **10,378** |

95% Confidence Intervals (CIs) in parentheses; DD-Dietary diversity; HH-Household; BMI-Body mass index
\*\*\* p<0.01, \*\* p<0.05, \* p<0.1

## Discussion

The study examined the socio-demographic factors associated with better child growth in poor households in five sub-Saharan African countries. We utilised a *positive deviance* approach as our analytical lens, whereby children who were not stunted though living in poor households were considered *positive deviants*. The findings showed that the effects of socio-demographic factors on child growth vary across countries. Maternal higher years of education had a significant positive effect on better linear growth among children living in poverty in DRC, Ghana, Kenya and Nigeria. This finding suggests that maternal education could mitigate the adverse effects of poverty on children’s nutritional status. Thus, education is an essential resource for improving child growth outcomes in the face of adversity. It is possibly the case because our conceptual framework (51) suggests that an exogenous variable such as education has a direct effect on caring practices, adequate dietary intake, utilisation of health care service and healthy environment among others. The proximate factors mentioned above, in turn, have direct effects on positive child growth outcomes. The literature on the effects of mothers education on child-caring practices and the utilisation of health services and the consequential positive effect on child health outcomes abound (57-60). Our study findings are consistent with the existing literature (59, 61). A study using data from three SSA countries showed that a higher level of maternal education was associated with reduced odds of child stunting (61). The literature, together with the present study, though using slightly different analytical approaches, demonstrated the importance of education in improving child growth outcomes.

Further, the results showed that in DRC and Ghana, mothers who attended at least four antenatal visits (ANC) have children with better linear growth outcomes. This could be due to the fact that mothers who attend ANC are likely to receive health and nutrition education, which may have a positive impact on their caring practices, with its consequential effect on better child health outcomes. The preceding explanation is in line with the conceptual framework used in this study, that posits that the proximate factors are pathways through which the exogenous factors influence child growth outcomes (51). Our findings are similar to others by previous researchers. For example, Kuhnt and Vollmer (62) found that having at least four ANC visits was associated with a reduced risk of stunting in pre-school children. Our findings, together with the literature, suggest that promoting ANC attendance among women can have a beneficial effect not only on the mothers but also their offspring. Therefore, interventions to promote child growth in poor environments should incorporate ANC as a critical intervention package.

Our analysis also illuminated the widely recognised benefits of breastfeeding for improved child health and developmental outcomes (63-65), but only when child-level covariates were not included in the empirical regression models. For instance, in Ghana, Mozambique and DRC, breastfeeding showed a significant positive effect on better childhood growth in the models containing only the socio-demographic factors. However, this significant association disappeared after child-level covariates such as dietary diversity, age and sex were included. This finding may mean that whether breastfeeding will have a positive effect on child growth or not depends to some extent on the inclusion or otherwise of child-level covariates. Therefore, in examining the effects of socio-demographic factors on child linear growth, it is significant to include child-level covariates to avoid presenting misleading estimates (66). The non-significant association between breastfeeding on child growth has previously been documented (60, 66, 67). Indeed, Marquis and colleagues (66) observed an inverse relationship between breastfeeding and child linear growth. They attributed this inverse relationship to what they termed *reverse causality*—that is, the breastfeeding did not lead to poor growth, but poor growth and health led to increased breastfeeding.

Surprisingly, in Mozambique, the widely recognised urban advantage in terms of favourable health outcomes was not observed in the present study. The analysis showed that urban place of residence associated negatively with child linear growth in poor households. The reason for this inverse relationship could be attributed to the precarious conditions under which some of the urban poor live (68). In the literature, both negative and positive effects have been found with the urban place of residence and child growth outcomes (68, 69). Some previous studies have observed that urban children are usually taller and heavier (69, 70). However, this may not include those children in poor urban settings, as there is evidence that children in these settings tend to have shorter heights than expected (68). This finding may mean that the so-called urban advantage does not benefit the urban poor in Mozambique.

### Strengths and limitations of the study

The use of large nationally representative samples provides more robust estimates of observed associations as well as enhancing the generalisability of the findings. The analysis of multi-country data helps to illuminate differences and highlights commonalities in the effects of the determinants on child growth across countries. The outcome variable was objectively measured, thereby reducing potential biases. The novelty of this analysis is the focus on factors that promote better child growth rather than risks factors for child growth deficiencies. A limitation worth mentioning is the cross-sectional nature of the data, which does not lend itself to the establishments of a causal relationship between the predictor and outcome variables. The conclusions in the paper are, therefore interpreted as mere associations between the predictor variables and the outcome variable. The use of quantitative data to investigate the PD approach may be a bit limiting as it may not be possible to explore all PD behaviours quantitatively. This limitation Notwithstanding, PD is a well-established concept and hence makes it possible to explore the approach (PD) using quantitative data.

## Conclusions

The study examined the effects of child, maternal, household and community-level sociodemographic factors on better linear growth among children in five SSA countries. The results showed that the effects of socio-demographic factors on child linear growth vary across countries. Maternal education has a positive effect on better growth among children in all countries except Mozambique. Improving maternal education in poor households may have a beneficial effect on child growth outcomes. A higher number of ANC visits has a significant positive effect on better child growth. Interventions to promote linear growth among children living in poverty should incorporate ANC as one of the critical intervention packages.

## Data Availability

This study was a re-analysis of existing data that are publicly available from The DHS Program at http://dhsprogram.com/publications/publication-fr221-dhs-final-reports.cfm. Data are accessible free of charge upon a registration with the Demographic and Health Survey program (The DHS Program). The registration is done on the DHS website indicated above.

## Declarations

### Consent for publication

The study used completely anonymous secondary data in the analysis. Therefore, no consent for publication was required

### Availability of data and materials

This study was a re-analysis of existing data that are publicly available from The DHS Program at http://dhsprogram.com/publications/publication-fr221-dhs-final-reports.cfm. Data are accessible free of charge upon registration with the Demographic and Health Survey program (The DHS Program). The registration is done on the DHS website indicated above.

### Authors’ Contribution

DAA conceived and designed the study, interpreted the results, wrote the first draft of the manuscript, and contributed to revision of the manuscript. DAA and ZTD analysed the data. ZTD contributed to study design, data interpretation, and critical revision of the manuscript. All authors take responsibility for any issues that might arise from the publication of this manuscript.

## Acknowledgements

We wish to express our profound gratitude to The DHS Program, USA for providing us access to the data. We also wish to acknowledge institutions of respective countries that played critical roles in the data collection process.

